# Modelling the Test, Trace and Quarantine Strategy to Control the COVID-19 Epidemic in the State of São Paulo, Brazil

**DOI:** 10.1101/2020.12.02.20242743

**Authors:** Marcos Amaku, Dimas Tadeu Covas, Francisco Antonio Bezerra Coutinho, Raymundo Soares Azevedo Neto, Claudio Struchiner, Annelies Wilder-Smith, Eduardo Massad

## Abstract

Testing for detecting the infection by SARS-CoV-2 is the bridge between the lockdown and the opening of society. In this paper we modelled and simulated a test-trace-and-quarantine strategy to control the COVID-19 outbreak in the State of São Paulo, Brasil. The State of São Paulo failed to adopt an effective social distancing strategy, reaching at most 59% in late March and started to relax the measures in late June, dropping to 41% in 08 August. Therefore, São Paulo relies heavily on a massive testing strategy in the attempt to control the epidemic.

Two alternative strategies combined with economic evaluations were simulated. One strategy included indiscriminately testing the entire population of the State, reaching more than 40 million people at a maximum cost of 2.25 billion USD, that would reduce the total number of cases by the end of 2020 by 90%. The second strategy investigated testing only symptomatic cases and their immediate contacts – this strategy reached a maximum cost of 150 million USD but also reduced the number of cases by 90%.

The conclusion is that if the State of São Paulo had decided to adopt the simulated strategy on April the 1^st^, it would have been possible to reduce the total number of cases by 90% at a cost of 2.25 billion US dollars for the indiscriminate strategy but at a much smaller cost of 125 million US dollars for the selective testing of symptomatic cases and their contacts.

## Introduction

Despite lockdowns in China, SARS-CoV-2 spread rapidly throughout the world by predicted routes of transportation (Bogoch et al., 2020) and spread faster than any other emerging infectious disease in recent decades (Angelo et al., 2019; Halstead & Wilder-Smith, 2019; Tuite et al., 2019; Wilder-Smith, Chang, & Leong, 2019). The first case of COVID-19 in Latin America was confirmed on 26 February 2020, in the São Paulo metropolis, the most populous city in the Southern hemisphere (∼12 million people) (Instituto Brasileiro de Geografia e Estatística, 2020). Travel reports and subsequent genetic analyses confirmed that the first detected infection was acquired via importation of the virus from Northern Italy (Candido et al., 2020; Jesus et al., 2020). Since then, Brazil has reported the largest number of cases in Latin America (4,238,446 reported cases and 129,522 deaths as of 11 September 2020) (Ministério da Saúde do Brasil, 2020). SARS-CoV-2 spread rapidly within Brazil (Carmo et al., 2020), often associated with urban centers with social deprivation (Souza et al., 2020) and has now been detected in the majority of the 27 federal states of Brazil. Using transmission pairs of SARS-CoV-2 reported to the Brazilian Ministry of Health, the mean and standard deviation for the serial interval was estimated to be 2.97 and 3.29 days respectively (Prete et al., 2020).

Social distancing, stay-at-home policies and discontinuation of mass gatherings up to complete lockdowns are important tenets of public health measures to mitigate the explosive growth of COVID-19 (Wilder-Smith & Freedman, 2020). However, the most effective public health measures include liberal testing, prompt isolation of all test positive persons, contact tracing of all test positive cases and enforced quarantining of all contacts (Hellewell et al., 2020; Salathe et al., 2020).

Here we set out to model the number of infections and deaths depending on the timing and extent of testing and contact tracing in the State of Sao Paulo, the most populous state of Brazil with 44,639,899 inhabitants (Biblioteca Virtual do Estado de São Paulo, 2020).

### The Model

The model is a modified version of the classical SEIR type of models (Massad et al., 2020) and considers that the population is divided into several compartments, namely: susceptible individuals at time *t, S*(*t*); tested susceptible individuals, *S*_*T*_ (*t*); exposed individuals, *E*(*t*); asymptomatic/oligosymptomatic individuals, *A*(*t*); infectious individuals, *I*(*t*); isolated infected individuals, *Q*(*t*); hospitalized individuals, *H*(*t*); individuals with severe disease hospitalized in intensive care units (ICU), *G*(*t*); and recovered individuals, *R*(*t*).

i. Susceptible individuals, denoted *S*(*t*), grow in number with a birth rate, and can either die by natural causes with rate *μ*, or acquire the virus with contact rate *β*, or be tested for the SARS-CoV-2, with rate *ξ*_*S*_ that begins at a time t_i_ after the begining of the outbreak.
ii. Tested susceptible individuals, denoted *S*_*T*_ (*t*), die by natural causes with the same rate *μ*, or acquire the virus with the same contact rate *β*.
iii. Once infected, the susceptible move to the state of Exposed, denoted *E*(*t*). These individuals either die by natural causes with the same rate *μ*, or evolve to the infectious individuals, denoted *I*(*t*), with rate *δ*_*I*_, or beggining at ime t_i_, be tested for the SARS-CoV-2, with rate *ξ*_*E*_ or evolve to asymptomatic/oligosymptomatic individuals, denoted *A*(*t*), with rate *δ*_*A*_.
iv. Infectious individuals, *I*(*t*), evolve for one of two states: Hospitalized individuals denoted *H*(*t*), or to a state in which individuals evolve to and develop severe disease, necessitating respiratory assistance, denoted *G*(*t*), with rates *σ*_*H*_ and *σ*_*G*_, respectively. Infectious individuals, *I*(*t*), also die by natural causes with rate *μ*, or by the disease, with rate *α*_*I*_, and can be tested tested for the SARS-CoV-2, with rate *ξ*_*I*_ that begins at time t_i_.
v. Individuals in the three states, *A*(*t*), *H*(*t*), and *G*(*t*), can die by natural causes with rate *μ*, or by the disease, with rates *α*_*A*_, *α*_*H*_, and *α*_*G*_, respectively. All indivuduals who acquired the infection and who did not die by the disease recover to a new state, denoted *R*(*t*), with rates *γ*_*I*_, *γ*_*A*_, *γ*_*H*_ and *γ*_*G*_, respectively.
vi. Tested individuals who resulted positive for the presence of the virus are quarantined in a stated denoted *Q*(*t*). Since these individuals are isolated from the rest of the population, they do not transmit the virus and will eventually recover from the infection, with rate *γ*_*Q*_, or will die by natural causes, with rate *μ*. A small fraction of asymptomatic individuals develop symptoms and are hospitalized with rate *σ*_*A*_.
vii. We assumed that the population birth rate Λ(*t*) is equal to the natural mortality of the population, not taking into account the disease-induced mortality.
viii. The fractions *p*_*E*_, *p*_*I*_, *p*_*A*_, *p*_*H*_, and *p*_*G*_ of exposed, symptomatic, asymptomatic, hospitalized and severe (ICU patients) individuals can transmit the infection.

The model’ states are shown in Figure 1.

**Figure 1.**
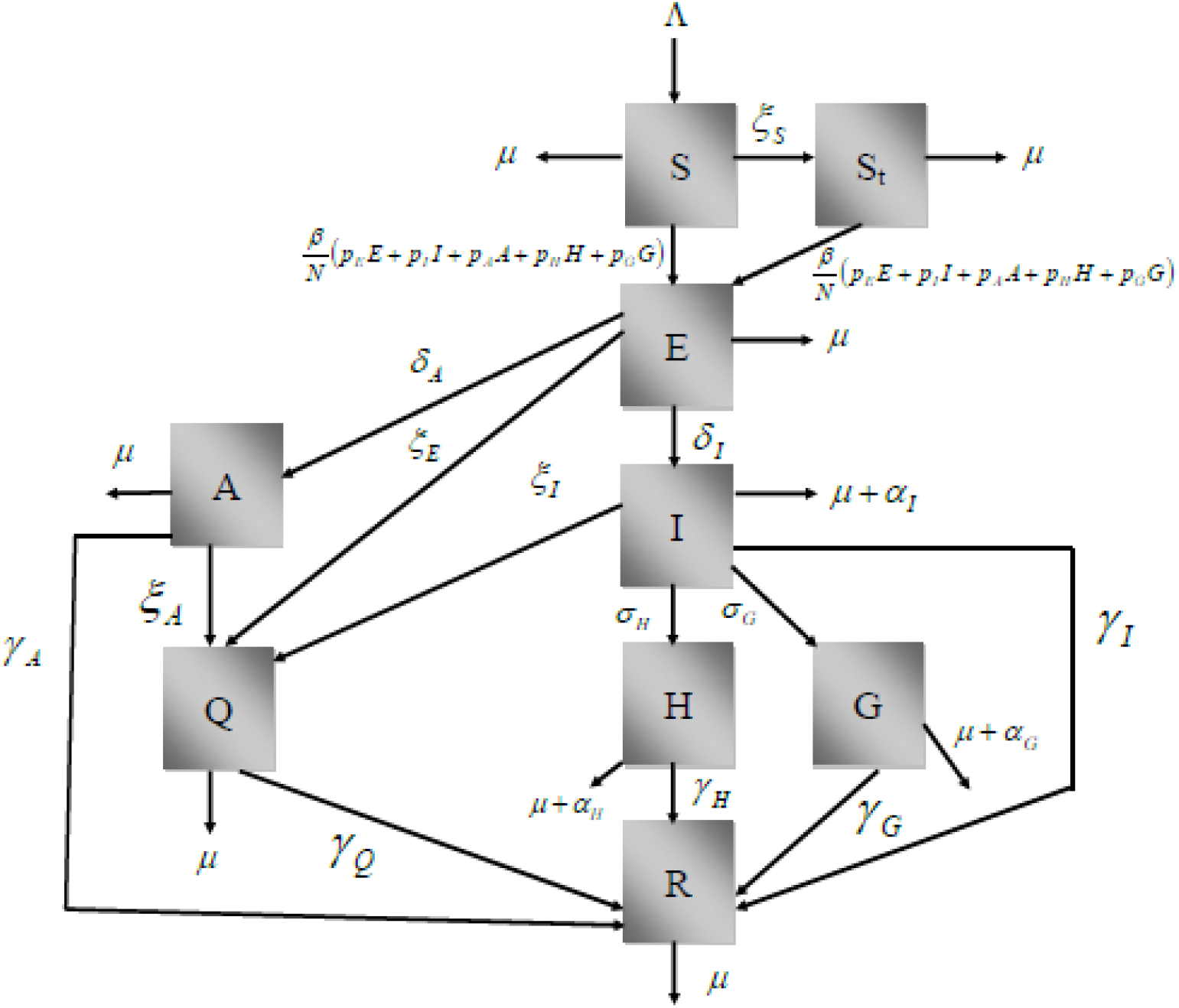
Schematic representation of the model compartments.

The model’s dynamics is described by the following set of differential equations:

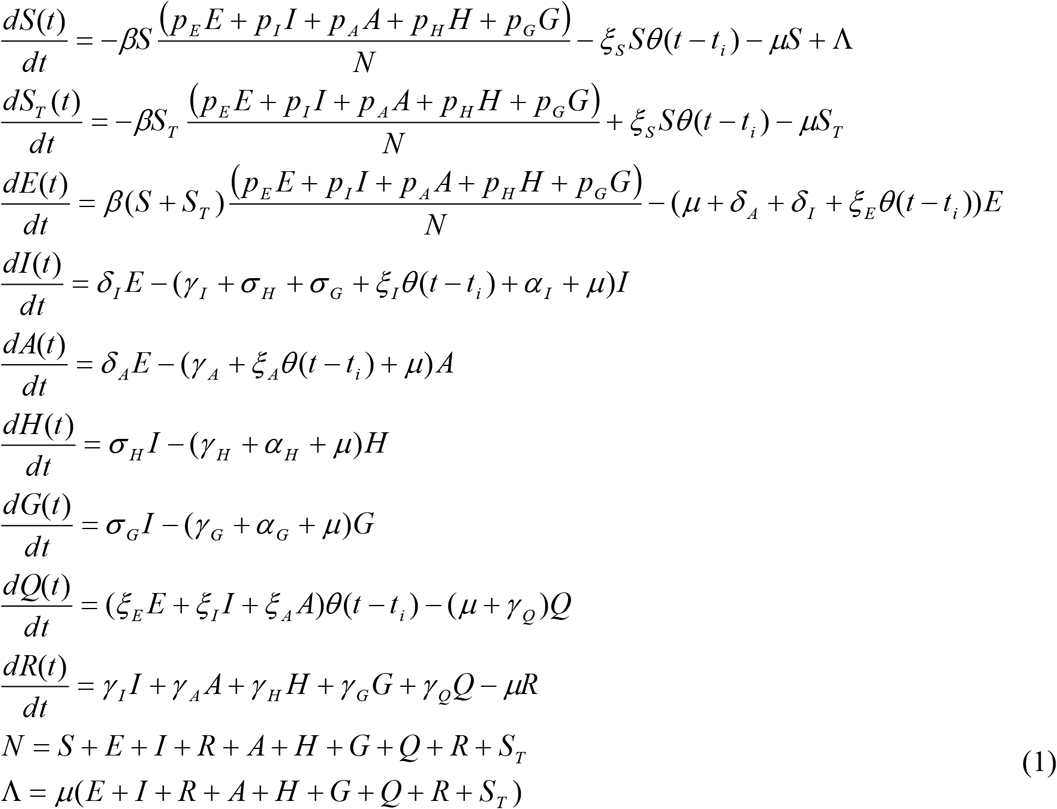

The Basic Reproduction Number of system (1) is given by:

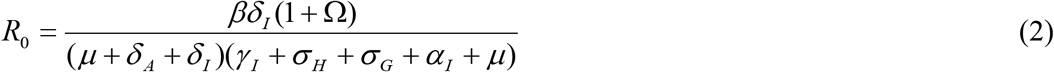

where:

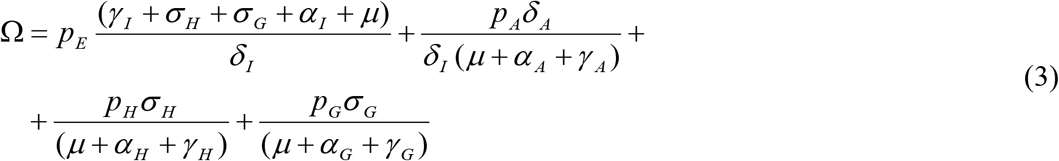

The incidence of infection is given by:

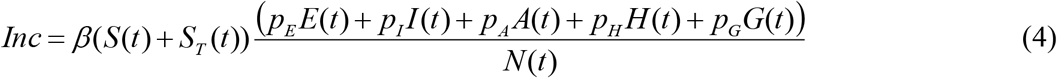

The total number of reported cases is obtained by multiplying the number of infected individuals by a notification ratio *K*(*t*):

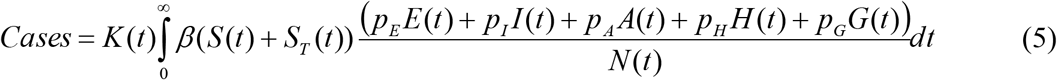

The total number of COVID-19-related deaths is given by:

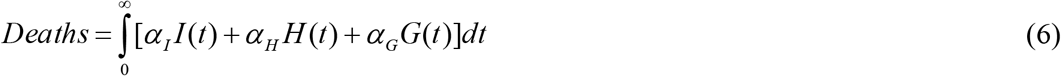

Finally, the total number of tested individuals is given by:

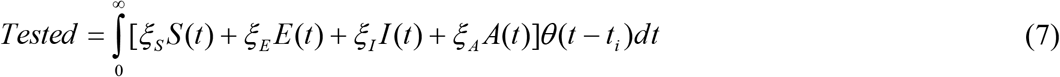

### Fitting procedure

Data on the cumulative number of reported cases and deaths were obtained from Seade (Fundação Sistema Estadual de Análise de Dados do Estado de São Paulo). Data on the number of ICU patients were obtained from SIMI (Sistema de Monitoramento Inteligente do Estado de São Paulo). A fitting procedure based on the Levenberg-Marquardt non-linear least-squares algorithm was used to fit the model’s parameters simultaneously to the data on cases, deaths and ICU patients. We used the R package minpack.lm (Elzhov et al., 2016).

We assumed that the potentially infective contact rate, the notification ratio, and the ICU admission rate change every 10 days. (As it will be explained in another paper this simulates the propagation of the disease and compliance with social distancing rules). The parameter values used are shown in Table 1.

**Table 1.** Parameters used in the model.

| Parameter | Description | Value |
| --- | --- | --- |
| $\beta(t)$ | Potentially infective contact rate | Fitted (changes over time) |
| $p_E$ | Infectivity of exposed individuals | 0.4* |
| $p_I$ | Infectivity of symptomatic individuals | 1.0* |
| $p_A$ | Infectivity of asymptomatic individuals | 1/3* |
| $p_H$ | Infectivity of hospitalized individuals | 0.01* |
| $p_G$ | Infectivity of ICU patients | 0.01* |
| $\mu$ | Natural mortality rate (life expectancy of 70 years) | $3.91 \times 10^{-5} \text{ days}^{-1}$ * |
| $\delta_I$ | Rate of evolution from exposed to infected | $1/2 \text{ day}^{-1}$ * |
| $\delta_A$ | Rate of evolution from exposed to asymptomatic | $1.45 \text{ day}^{-1}$ ** |
| $\gamma_I$ | Rate of recovery from infected | $1/3 \text{ day}^{-1}$ * |
| $\gamma_A$ | Rate of recovery from asymptomatic | $1/14 \text{ day}^{-1}$ * |
| $\gamma_H$ | Rate of recovery from hospitalized | $1/10 \text{ day}^{-1}$ * |
| $\gamma_G$ | Rate of recovery from ICU | $0.06752 \text{ day}^{-1}$ ** |
| $\gamma_Q$ | Rate of recovery from isolated | $1/14 \text{ day}^{-1}$ * |
| $\alpha_I$ | Disease-induced mortality rate for infected individuals | $5 \times 10^{-4} \text{ day}^{-1}$ * |
| $\alpha_A$ | Disease-induced mortality rate for asymptomatic individuals | 0 * |
| $\alpha_H$ | Disease-induced mortality rate for hospitalized individuals | $2.2012 \times 10^{-4} \text{ day}^{-1}$ ** |
| $\alpha_G$ | Disease-induced mortality rate for ICU patients | Fitted (changes over time) |
| $\xi_S$ | Testing rate of susceptible individuals | Variable |
| $\xi_E$ | Testing rate of exposed individuals | Variable |
| $\xi_I$ | Testing rate of symptomatic individuals | Variable |
| $\xi_A$ | Testing rate of asymptomatic individuals | Variable |
| $\sigma_H$ | Hospitalization rate | $1.973 \times 10^{-2} \text{ day}^{-1}$ ** |
| $\sigma_G$ | ICU admission rate | Fitted (changes over time) |
| $K(t)$ | Notification ratio | Fitted (changes over time) |
| $\Lambda(t)$ | Birth rate | Changes over time |

\*assumed; \*\*fitted

Model projections for future dates were obtained by keeping fixed the fitted values of the parameters from the last date observed in the data.

### The Test-Trace-Quarantine strategy

Testing, tracing and quarantining positive individuals simulated consists in applying 10 thousand health agents from the programme of primary health of the State Secretary of Health that would visit the 3,933,448 dwellings of the State of São Paulo. In each dwelling, the agent would interview the inhabitants questioning who had had any of the symptoms of COVID-19 since March 2020 and about who had had contact to any individual who had the disease in the same period. In case of any positive answer the individuals would be tested for the presence of the virus with the RT-PCR technique. In case of positive result, the individual would be quarantined for 14 days.

The cost of the strategy was calculated assuming the cost of the test, assumed the price tag of US$48.00 per sample and the cost of the agenst salary, assumed to be US$1,128.00 per month. The total cost of each simulated strategy would depend on the rate and of the total number of testing, as well as the duration of each strategy.

## Results

We fitted the model’s parameters simultaneously to the data of cumulative number of reported cases and deaths (Figures 2(a) and 2(b), respectively), and the number of ICU patients for the state of São Paulo until July 18, 2020. To estimate a 95% probability interval (shaded area in Figure 2), we assumed a normal distribution for the contact rate with a standard deviation of 1.5%. The fitted parameters are shown in Table 1.

**Figure 2.**
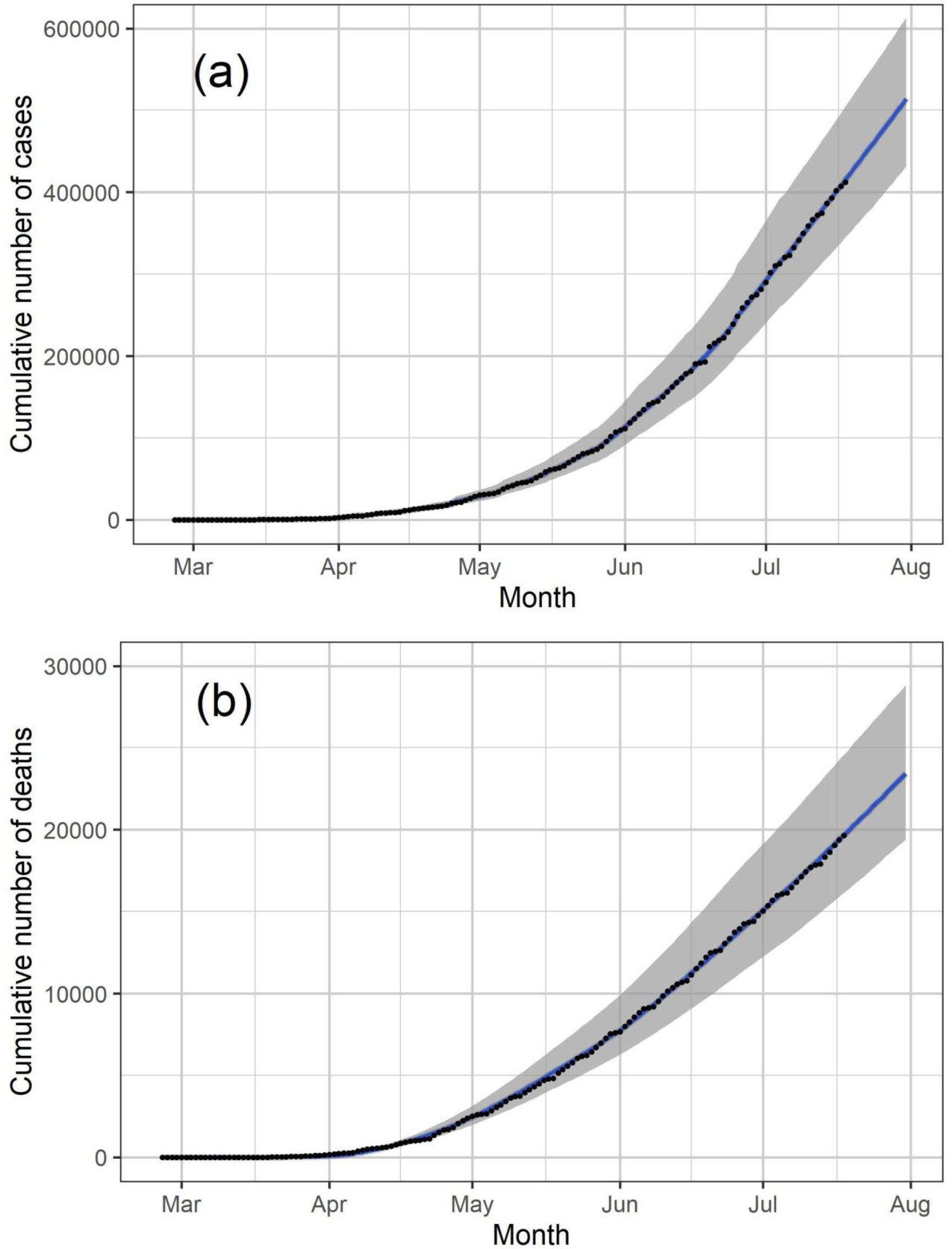
Cumulative number of reported cases and deaths (black dots in (a) and (b), respectively) and the corresponding fitted model (blue lines). The solid lines and shaded area correspond, respectively, to median values and 95% probability intervals.

The results for the strategy that considers testing susceptible and infected (symptomatic and asymptomatic) individuals are shown in Figure 3. We calculated a testing strategy efficacy, subtracting from 1 the result of the division of the number of cumulative cases under a specific testing strategy up to December 31, 2020 by the number of reported cases in a scenario in which no test is used. The testing strategy efficacies as a function of the total number of tests and corresponding costs in US dollars are shown in Figures 3(a) and 3(b), respectively, for different times t_i_, the strategy started to be applied. We noticed that a 50% reduction in the number of cases is only achieved when the total number of tests is approximately 40 million (the state population is about 44.6 million people) with an approximate total cost of 2 billion US dollars, except for the start date of August 1^st^.

**Figure 3.**
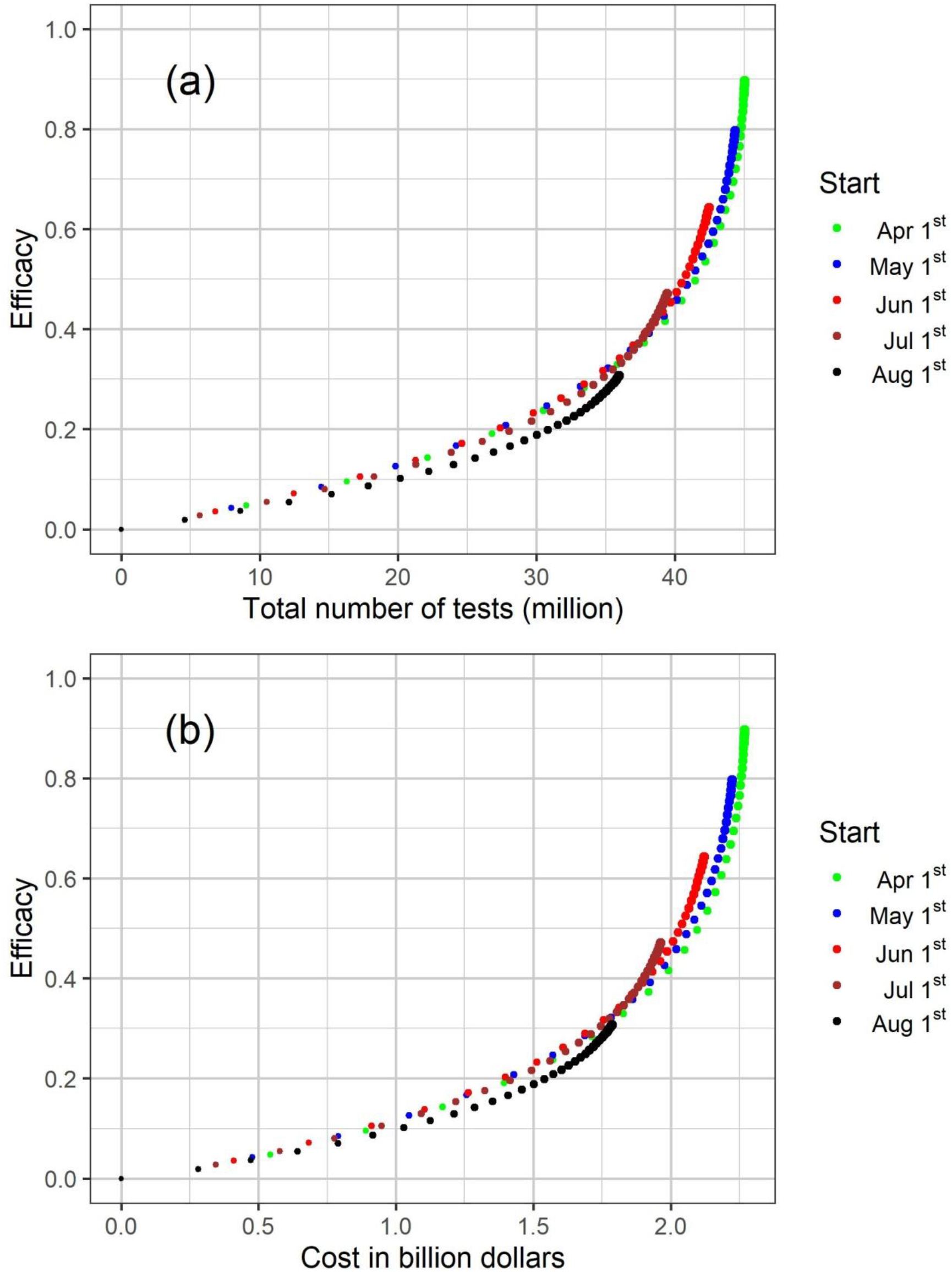
Results for the strategy that considers testing susceptible and infected (symptomatic and asymptomatic) individuals showing the evolution of the testing strategy efficacy (1 minus the number of cumulative cases under a specific testing strategy up to December 31, 2020 divided by the number of cases when no test is used) as a function of the total number of tests (a) and corresponding costs in US dollars (b) for different dates of start. Each dot corresponds to a different daily testing rate and the dot size is proportional to the testing rate.

The results for the strategy that considers testing infected (symptomatic and asymptomatic/oligosymptomatic) individuals are shown in Figure 4. The testing strategy efficacies as a function of the total number of tests and corresponding costs in US dollars are shown in Figures 4(a) and 4(b), respectively, for different dates of start.

**Figure 4.**
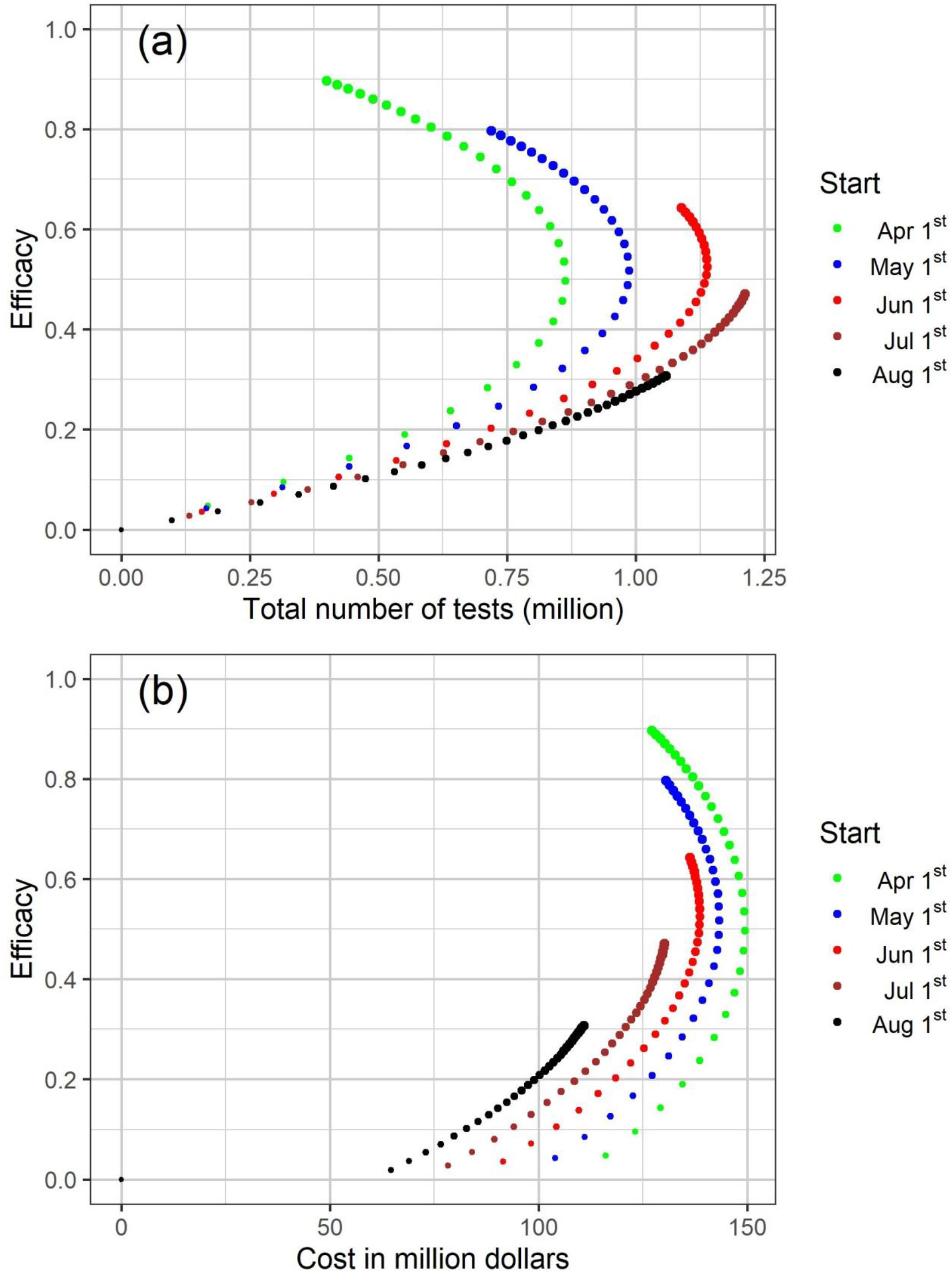
Results for the strategy that considers testing infected (symptomatic and asymptomatic) individuals showing the evolution of the testing strategy efficacy (1 minus the number of cumulative cases under a specific testing strategy up to December 31, 2020 divided by the number of cases when no test is used) as a function of the total number of tests (a) and corresponding costs in US dollars (b) for different dates of start. Each dot corresponds to a different daily testing rate and the dot size is proportional to the testing rate.

In Figures 3 and 4, each dot corresponds to a different daily testing rate and the dot size is proportional to the testing rate. Dot sizes and corresponding testing rates can be observed in Figure 5, in which we observe that higher testing rates and earlier start dates are more efficacious.

**Figure 5.**
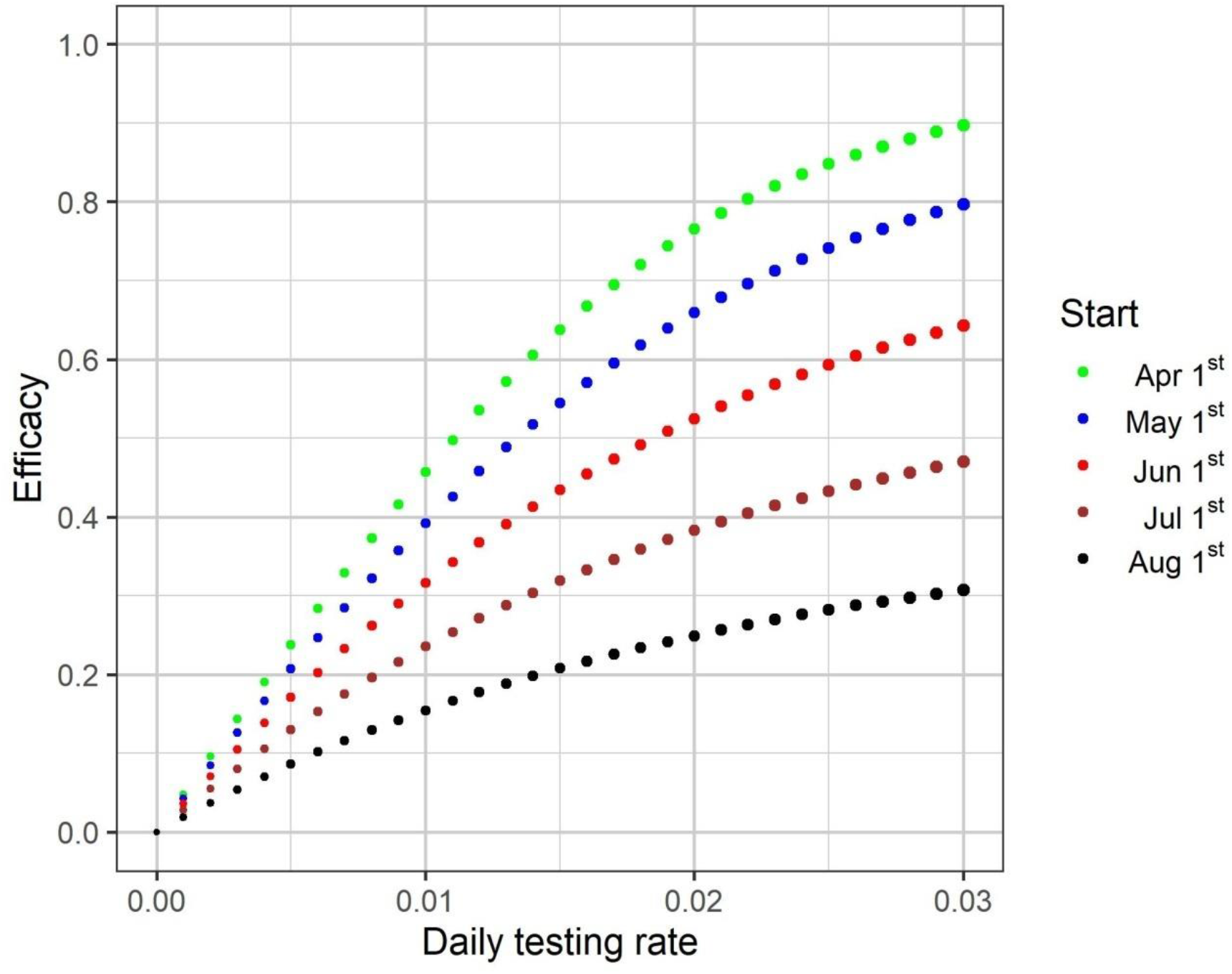
Results for the strategy that considers testing infected (symptomatic and asymptomatic) individuals showing the evolution of the testing strategy efficacy (1 minus the number of cumulative cases under a specific testing strategy up to December 31, 2020 divided by the number of cases when no test is used) as a function of the daily testing rate for different dates of start. The dot size is proportional to the testing rate.

In Figure 4(a), dots in the upper part of the graph correspond to higher testing rates. For each date of start, we noticed that dots corresponding to different testing rates are displayed in a concave shape. Thus, depending on the testing rate, similar total number of tests may result in different efficacies. Let us consider the specific case of the testing rates 0.5% and 2.1% per day for the start date of April 1^st^ (Figure 6). We noticed in Figure 4(a) that the total number of tests used until December 31, 2020, is similar (approximately 0.63 million tests) but the cumulative numbers of cases are about 0.95 and 0.27 million cases for the rates 0.5% and 2.1% per day, respectively. In the model, the number of tested individuals is proportional to the number of infected people. For a low testing rate, the initial number of individuals tested and consequently the number of individuals in quarantine are both small, and the force of infection and thus the disease spread are not efficiently reduced. For a high testing rate, on the other hand, the initial number of individuals tested and consequently the number of individuals in quarantine are larger, hence the force of infection and the number of new cases are both reduced more efficiently over time.

**Figure 6.**
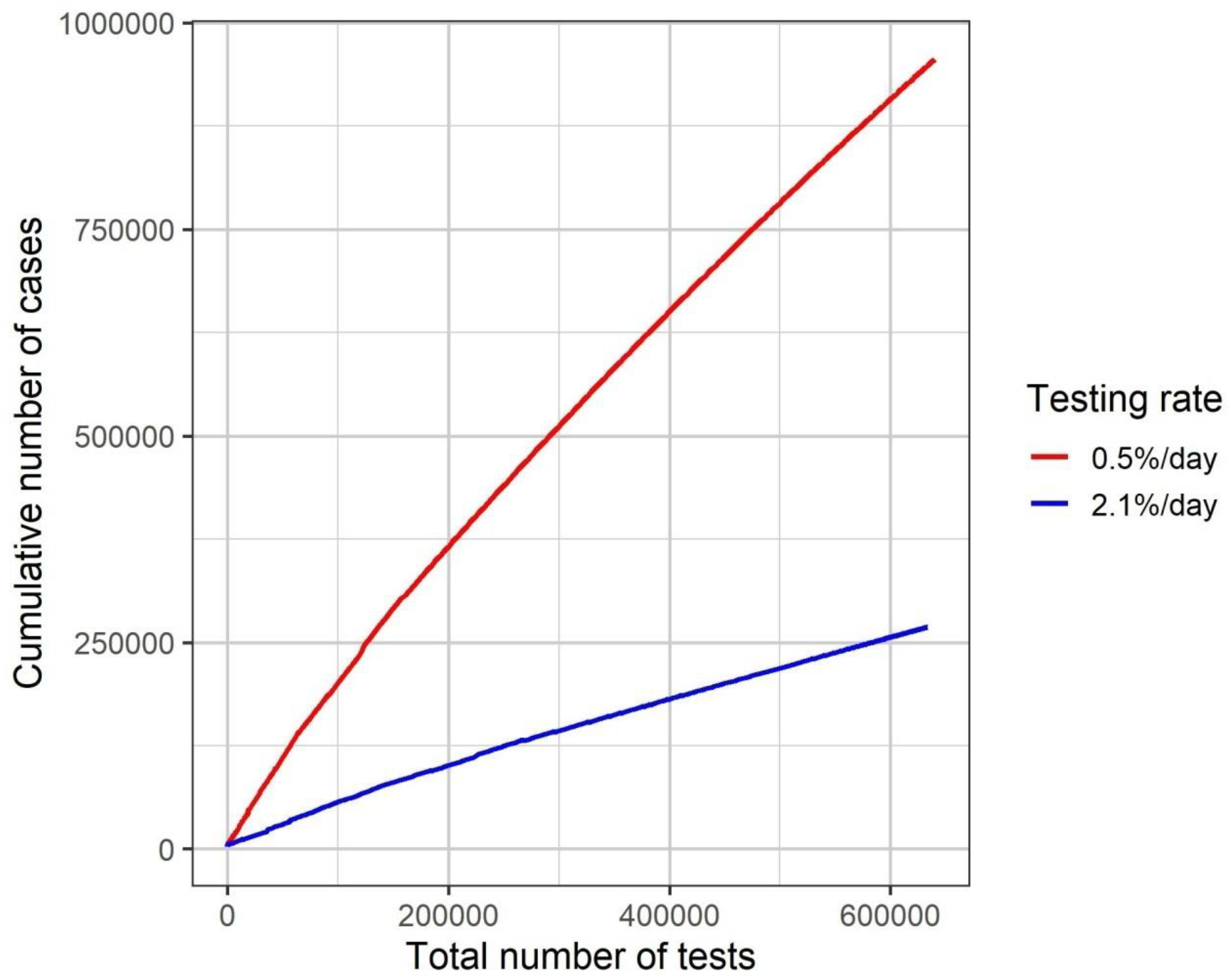
Total number of tests over time and the corresponding number of cumulative cases for two different daily testing rates (0.5% and 2.1% per day) for mass testing starting on April 1^st^.

When only (suspected) infected individuals are tested, the total cost is higher for earlier start dates (Figure 4(b)), because operational costs are substantionally higher than the costs related to the diagnostic tests.

## Discussion

We modelled and simulated a test-trace-and-quarantine strategy to control the COVID-19 outbreak in the State of São Paulo, Brasil. The State of São Paulo failed to adopt an effective social distancing strategy, reaching at most 59% in late March and started to relax the measures in late June, dropping to 42% in 11 September (Sistema de Monitoramento Inteligente do Estado de São Paulo, 2020). Therefore, São Paulo relies heavily on a massive testing strategy in the attempt to control the epidemic.

We simulated two alternative strategies combined with economic evaluations. One strategy included indiscriminately testing the entire population of the State, reaching more than 40 million people at a maximum cost of 2.25 billion USD, that would reduce the total number of cases by the end of 2020 by 90%. The second strategy investigated testing only symptomatic cases and their immediate contacts – this strategy reached a maximum cost of 150 million USD but also reduced the number of cases by 90%. As we were interested in the simulation of the impact of testing and quarantining in the spread of the infection in the general population, we did not included testing for other clustering. For instance, health workers are tested regularly in Sao Paulo and are isolated when tested positive.

Our model has the following limitations. The model assumes a perfect test with 100% sensitiviy and specificity; we also assumed that tracing the contacts of the positive cases would be perfect, which is obviously a practical impossibility. Another important limitation of the model is that it does not consider any delay in the results of the test, an important factor that has been limiting this strategy in many places of the world (Kretzschmar et al., 2020). Although the current tests have a delay of many days to present the results, many rapid-molecular point of care tests for detecting positive patients to SARS-CoV-2 are in development (Ahmad et al., 2020), so the delay in test results can be overcome. However, our model does not take into account social inequity in accessing testing (Souza, Carmo, & Machado, 2020; Martins-Filho et al., 2020). Some strategies could be considered to reduce the testing costs like pooling samples for testing (Abid et al., 2020) or concentrating the tests in neighborhood where sewege systems were tested positive (Foladori et al., 2020).

## Conclusion

Had the State of São Paulo decided to adopt this strategy on April the 1^st^, it would have been possible to reduce the total number of cases in 90% at a cost of 2.25 billion US dollars for the indiscriminate strategy but at a much smaller cost of 125 million US dollars for the selective testing of symptomatic cases and their contacts. We conclude that a selective test, trace and quarantine strategy is the most cost effective strategy that could be applied in situations where social distancing is difficult to implement.

## Data Availability

The data on COVID-19 cases and deaths are available in https://www.seade.gov.br/

## Authors contributions

All authors contributed equally in all phases of this work.

### Acknowledgements

This work was partially supported by the project ZikaPLAN, funded by the European Union’s Horizon 2020 research and innovation programme under Grant Agreement No. 734584, by LIM01-HFMUSP, CNPq and FAPESP and Fundacao Butantan.

## Ethical approval

Not applicable (this is a purely theoretical work with no human subject involved).

